# The association of pregnancy with disease progression in patients previously treated for differentiated thyroid cancer: A propensity score-matched retrospective cohort study

**DOI:** 10.1101/2023.03.15.23287341

**Authors:** Xin Li, Wu-Cai Xiao, Fang Mei, Rui Shan, Shi-Bing Song, Bang-Kai Sun, He-Ling Bao, Jing Chen, Chun-Hui Yuan, Zheng Liu

## Abstract

**IMPORTANCE:** Differentiated thyroid cancer (DTC) is increasingly common in women of reproductive age. However, whether pregnancy increases the risk of progression/recurrence of DTC after treatment remains controversial due to the effect of confounding.

**OBJECTIVE:** To assess the effect of pregnancy on structural or biochemical progression in patients previously treated for DTC in a retrospective cohort using propensity score matching (PSM).

**DESIGN, SETTING, AND PARTICIPANTS:** This cohort study included 123 pregnant women and 1,376 non-pregnant women after initial treatment for DTC at Peking University Third Hospital between January 2012 and December 2022. To control the effect of confounding, we carefully matched pregnancy (n = 102) and non-pregnancy groups (n = 297) in terms of age, Hashimoto’s thyroiditis, lymph node dissection, extra-thyroid invasion, initial risk of recurrence after treatment, and time interval between treatment and last follow up by using PSM.

**EXPOSURES:** DTC patients became pregnant after previous treatment.

**MAIN OUTCOMES AND MEASURES:** The risk of structural or biochemical progression was assessed in the pregnancy and PSM matched non-pregnancy groups, respectively.

Conditional logistics regression models were used to control important confounders and consider the matching properties of the data.

**RESULTS:** At baseline, the pregnancy (n = 102) and non-pregnancy groups (n = 297) were balanced in all matched variables (standardized differences <10% and *P* > 0.05). After a mean follow-up of approximately 4.5 years, we observed no evidence of difference between the two groups in growth in the size of existing metastatic foci [2 (2.0 %) vs. 2 (0.7 %); *P* = 0.346], percentage of patients developing new lymph node metastases [4 (3.9 %) vs. 21 (7.1 %); *P* = 0.519], node growth in the contralateral thyroid lobe [4 (3.9 %) vs. 16 (5.4 %); *P* = 0.324 ], or biochemical progression [2 (2.0 %) vs. 9 (3.0 %); *P* = 0.583]. Results from conditional logistic regressions and several sensitivity analyses also showed no evidence of association of pregnancy with the risk of progression, after adjusting for potential confounders of age, tumor size, initial risk stratification, Hashimoto’s thyroiditis, lymph node dissection, the time interval between treatment and follow-up, and achievement of TSH inhibition target (*P* = 0.354). The pregnancy-progression association observed longer than 4.5 years showed no evidence of difference with that observed shorter than 4.5 years (*P* for interaction was 0.283). We further classified the pregnancy patients into 3 subgroups based on the time interval between treatment and pregnancy (< 1 year, 1-2 years, ≥ 2 years) and found that the shorter the time interval, the higher the risk of DTC progression (*P* for trend was 0.043).

**CONCLUSIONS AND RELEVANCE:** The risk of DTC progression/recurrence in the pregnant women was not higher than that in the well-matched, non-pregnant women. For young women previously treated for DTC, disease progression might not be a concern for their future pregnancy plan, but it seems safer to wait an appropriate amount of time before pregnancy.

**Key Points:** *Question:* Does pregnancy increase the risk of disease progression/recurrence in patients previously treated for differentiated thyroid cancer (DTC)?

*Findings:* This propensity score-matched retrospective cohort study included 399 patients previously treated for DTC. In a mean follow-up of approximately 4.5 years, the risk of progression in the pregnant group was not higher than the well-matched, non-pregnancy group, but the shorter time interval between treatment and pregnancy (≤ 2 years) appeared to increase the risk of disease progression.

*Meaning:* For young women previously treated for DTC, disease progression might not be a concern for their future pregnancy plan, but it seems safer to wait an appropriate amount of time before pregnancy.

## Introduction

Globally, thyroid cancer has become the 5^th^ most commonly diagnosed cancer in adult women ^1^. In China, the female age-standardized incidence rates of thyroid cancer have substantially increased from 6.68/10^5^ in 2005 to 20.28/10^5^ in 2015 ^2^. Notably, the annual burden of new cases of thyroid cancer in the female population was approximately tripling that in the male population ^2^. Differentiated thyroid cancer (DTC), including papillary and follicular cancer, accounts for more than 90% of all thyroid cancers ^3^. DTC has been the 2^nd^ most common cancer only following breast cancer in women of reproductive age ^4^.

Young women diagnosed with DTC often had great concerns about their future pregnancy plans ^5^. The pregnancy process is accompanied by an increase of hormones of estrogen and human chorionic gonadotropin, which might play a role in the pathogenesis, progression/recurrence, and metastasis of thyroid tumors. Clinicians should be aware of the potential risk of pregnancy on DTC progression so they can counsel patients appropriately. DTC before pregnancy has been classified as moderate-to-high risk among all thyroid diseases before pregnancy, according to the most recent guidelines (2022) for the prevention and management of thyroid diseases during pregnancy and perinatal period in China ^6^. However, clinical controversy exists in the management of pregnancy-associated DTC, based on our systematic review of this topic^7^.

To date, the overall research evidence to clarify this research question has been scarce in both quantity and quality. Two studies^8,9^ used the single-group design and the study subjects were exclusively pregnant women. Of note, findings from this study design could not elucidate whether the disease progression of DTC was attributed to the pregnancy process per se or just a natural process of disease likely existing in both pregnancy and non-pregnancy patients. Only 3 studies from Morocco, China, and Japan have used the controlled-group design: comparing disease progression between the pregnancy and non-pregnancy patients previously treated for DTC ^10-12^. However, none of them have adjusted potential confounders associated with the progression of DTC such as Hashimoto’s thyroiditis ^13^. Moreover, the sample size was small, with the largest pregnancy group only including 42 patients. Additionally, a study has indicated that the interval between treatment and pregnancy might influence survival outcomes among patients with breast cancer, and patients who became pregnant 2 years after surgery had better survival outcomes than those who became pregnant immediately after surgery^14^. But to our knowledge, no studies have examined the effect of the interval between treatment and pregnancy on disease progression of DTC.

To address this, our study aimed to rigorously clarify the association of pregnancy with the disease progression of DTC by comparing the pregnancy group with the non-pregnancy group and using a propensity score matching (PSM) method, which could reduce the effect of confounding in observational studies^15^. We would also explore the role of the time interval between surgery and pregnancy in DTC progression. The findings of our study will provide the evidence required for counseling and management of pregnancy-associated DTC in clinical practice.

## Materials and Methods

### Study population

This was a retrospective cohort study. We reviewed medical records of patients with DTC who received treatment and follow-up examinations at Peking University Third Hospital from January 2012 to December 2022. We included patients who satisfied the following 4 criteria: (1) diagnosis of DTC, (2) treatment with total thyroidectomy or lobectomy, (3) records of birth delivery, and (4) follow-up examinations of neck ultrasonography, or measurements of serum Tg levels or Tg antibodies. Of the 1,505 patients satisfying these criteria, 129 become pregnant after treatment and 1,376 did not. Of the 129 pregnant women, 6 were excluded due to twin delivery (n = 2), preterm birth (n = 2), stillbirth (n = 1), or spontaneous abortion (n = 1) (Figure 1). Then, patients who became pregnant after treatment for DTC [for a median of 1.73 (range: 0.09-6.63) years] were included in the pregnancy group. Finally, a total of 123 and 1,376 patients were included in the pregnancy group and non-pregnancy group, respectively. This study was approved by the Medical Research Ethics Committee of Peking University Third Hospital (No. IRB00006761-M2022721).

**Figure 1.**
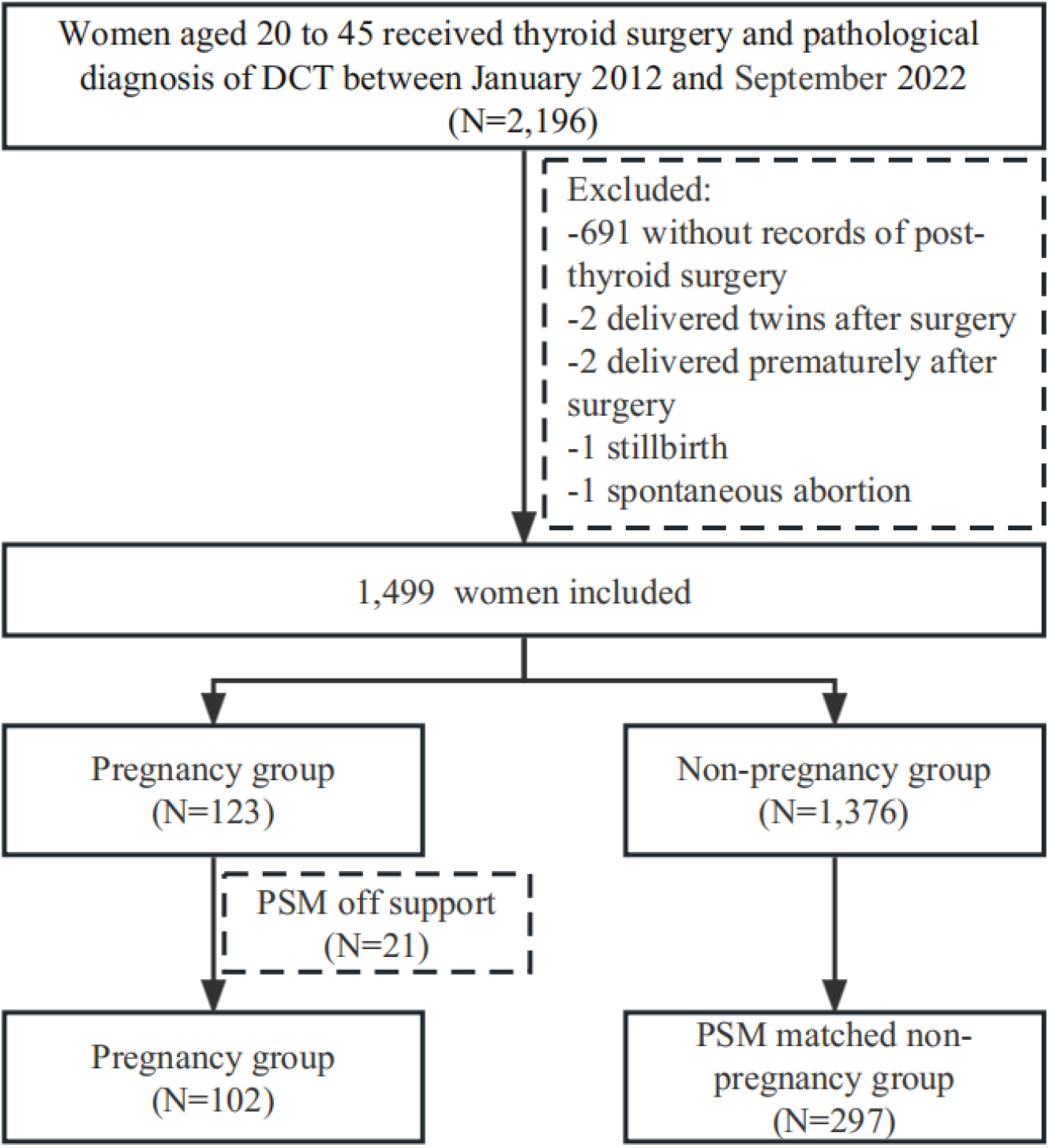
Flow chart of object inclusion, exclusion and propensity score matching

### Data extraction

Data were carefully extracted from clinical records by two researchers with rich experiences in clinical practice (XL) and data pre-processing (WCX). We extracted detailed information throughout the preoperative, surgical, postoperative, and post-delivery periods, including patients’ age, the severity of cancer (tumor size, lymph node metastasis, extra-thyroid invasion), surgery type, birth delivery (last menstrual period, delivery date, delivery outcomes), measurements of thyrotropin (TSH), Tg levels, and Tg antibodies, and the existence of Hashimoto’s thyroiditis.

Serum Tg levels were measured with thyroglobulin assay (Elecsys Tg II, Roche Diagnostics, Penzberg, Germany). Tg antibodies and TSH levels were measured with commercial kits (Siemens Healthcare Diagnostics) using a fully automatic chemiluminescence immunoassay analyzer (ADVIA Centaur XP, Siemens Healthcare Diagnostics).

### Risk stratification at baseline before pregnancy

To estimate risks of recurrence and disease-specific mortality in the study subjects at baseline, we performed risk stratification of patients by using three common approaches in the modern management of DTC ^16^. The first approach was to stratify patients into tumor-node-metastasis (TNM) stage according to the 8^th^ edition of the American Joint Committee on Cancer (AJCC) staging system, considering the factors of tumor size, extra-thyroid invasion, lymph node metastasis, and distant metastasis ^17^. In the second approach, patients were stratified into the high, intermediate, or low risk of recurrence according to the modified 2015 American Thyroid Association (ATA) risk stratification system ^18^. For patients stratified into high, intermediate, or low risk, TSH suppression was recommended to maintain below 0.1, within 0.1-0.5, or 0.5-2 mU/L, respectively. We also stratified patients in the pregnancy group into 4 categories by using the response-to-therapy assessments before pregnancy: excellent, indeterminate, biochemical incomplete, and structural incomplete response. For patients treated with lobectomy, response-to-therapy assessments were classified based on the definitions published before ^19^.

### Disease progression/recurrence at follow-up after delivery

We assessed the progression/recurrence of DTC at follow-up in both structural and biochemical types. Structural progression referred to ≥ 3 mm growth in the size of existing metastatic foci ^8^, the development of new lymph node metastases, or ≥ 2 mm growth in the size of existing cancer foci in the contralateral thyroid by using neck ultrasonography. Biochemical progression referred to a 20% or more increase in serum Tg or Tg antibodies relative to the pre-surgery level ^20^. We excluded patients if the presence of serum Tg antibodies precluded the accuracy of measurements of Tg levels.

### Statistical Analyses

We used propensity score matching to obtain matching cohorts from the included population. Firstly, we calculated propensity scores (PS) for pregnancy after surgery using logit regression. The variables included in the model were age at the time of surgery, presence or absence of Hashimoto’s thyroiditis, lymph node dissection at the time of surgery, external thyroid invasion, grade of risk of recurrence after surgery, and time interval between surgery and last follow up. Then, we performed a 1:4 matching on the logit of PS with a caliper of width equal to 0.2 of the standard deviation of the logit of the PS^21^. We used standardized differences to compare the balance of covariate distributions between the matched pregnancy and non-pregnancy groups, which took into account the matching properties of the sample^22,23^. A standard difference of less than 10% indicates a negligible difference in the mean or prevalence of covariates between matched groups^15^.

We compared baseline characteristics between pregnancy and original (unmatched) non-pregnancy groups by using the t-test and Kruskal-Wallis rank sum test for normally and abnormally distributed continuous variables, respectively, and using the Pearson chi-square test or Fisher’s exact probability test (when the number of cases was ≤ 5) for categorical variables. We also compared baseline characteristics between pregnancy and PSM matched non-pregnancy groups by using the t-test of regression based on grouping variables and weighted with matching weights. We used 3 conditional logistics regression models to assess the crude and adjusted associations of pregnancy with the progression risk of DTC after delivery, which could also take into account the matching properties of the data. In Model 1, we did not adjust any covariates (crude model); in Model 2, we adjusted preoperative tumor size, the presence of Hashimoto’s thyroiditis, lymph node dissection, and the time interval between surgery and follow-up; in Model 3, we additionally adjusted age, and achievement of TSH inhibition target based on Model 2. We divided subgroups according to whether the follow-up time was longer than the median follow-up time and analyzed differences in outcomes between subgroups. We also conducted sensitivity analyses by excluding 11 patients who were recommended for I^131^ radioactive iodine treatment. Statistical analyses were performed using R software version 4.2 and Stata software version 16.0. *P* values < 0.05 were considered statistically significant.

## Results

### Baseline characteristics of patients with DTC

The PS of 21 patients in the pregnancy group were not in the same range as those in the non-pregnancy group (off support), so they were not included in the analysis. The process of sample inclusion and matching can be seen in Figure 1.

We compared baseline characteristics between the pregnancy group (n = 102) and the unmatched/matched non-pregnancy groups (n = 1,376/297) in Table 1. The baseline characteristics between pregnant and non-pregnant groups were not balanced before matching. Specifically, compared with the original non-pregnant group, the pregnant group has a lower age, a lower proportion of primary tumour status of T3b and a lower recurrence after treatment (*P* < 0.001). As expected, patients in the pregnancy group and the matched non-pregnancy group were well comparable in age, TNM stage, and initial risk stratification after treatment, Hashimoto’s thyroiditis, extra-thyroid invasion, and lymph node dissection, except for surgery type (standardized differences <10% and *P* > 0.05).

**Table 1.** Baseline characteristics of pregnancy group, unmatched non-pregnancy group, and PSM matched non-pregnancy group

| Characteristics | Pregnancy group<br>(n=102) | Non-pregnancy group |  |  |  |
| --- | --- | --- | --- | --- | --- |
|  |  | Unmatched<br>(n=1376) | *P*<sup>1</sup> | *PSM matched*<br>(n=297) | *P*<sup>2</sup> |
| **Age at surgery (year)** | 30.54 ± 4.12 | 33.75 ± 6.61 | <0.001 | 31.00 ± 6.36 | 0.501 |
| **Primary tumor** |  |  |  |  |  |
| Maximum diameter of tumor (cm) | 1.19 ± 0.78 | 1.24 ± 0.83 | 0.584 | 1.22±0.80 | 0.763 |
| Primary tumor status, n (%) |  |  | <0.001 |  | 0.227 |
| T1a | 54(52.9) | 647 (47.0) |  | 167(56.2) |  |
| T1b | 38 (37.3) | 372(27.0) |  | 84 (28.3) |  |
| T2 | 6 (5.9) | 105(7.6) |  | 34(11.5) |  |
| T3a | 2(2.0) | 15 (1.0) |  | 3(1.0) |  |
| T3b | 2 (2.0) | 237(17.2) |  | 9(3.0) |  |
| **Nodal status at diagnosis, n (%)** |  |  | 0.652 |  | 0.231 |
| N0 | 84 (82.4) | 1131 (82.2) |  | 263(88.6) |  |
| N1a | 13(12.8) | 150 (10.9) |  | 22(7.4) |  |
| N1b | 5(4.9) | 295(6.9) |  | 12(4.0) |  |
| **Metastases at diagnosis** |  |  |  |  |  |
| M0 | 102 (100) | 1376 (100%) | >0.999 | 297(100) | >0.999 |
| **Recurrence risk after treatment, n (%)** |  |  | <0.001 |  | 0.949 |
| High | 2 (2.0) | 237(17.2) |  | 9 (3.0) |  |
| Intermediate | 6 (5.9) | 127 (9.2) |  | 17 (5.7) |  |
| Low | 94 (92.2) | 1012 (73.6) |  | 271(91.3) |  |
| **Hashimoto's thyroiditis, n (%)** | 39(38.2) | 602(43.8) | 0.278 | 114(38.4) | >0.999 |
| **Extra-thyroid invasion, n (%)** | 7(7.9) | 250(18.2) | 0.419 | 19 (6.4) | 0.720 |
| **Surgery type, n (%)** |  |  | 0.860 |  | 0.032 |
| Total thyroidectomy | 34 (33.3) | 447 (32.5) |  | 59 (19.9) |  |
| Lobectomy | 68(66.7) | 929 (67.5) |  | 238(80.1) |  |
| **Lymph node dissection, n (%)** | 18(17.7) | 242 (17.6) | 0.961 | 33(11.1) | 0.978 |

**Note:**<sup>1</sup>T-test and Kruskal-Wallis rank sum test for normally and abnormally distributed continuous variables, respectively, and using the Pearson chi-square test or Fisher's exact probability test (when the number of cases was ≤ 5) for categorical variables. <sup>2</sup>T-test of regression based on treatment variables and weighted with matching weights.

### Follow-up outcomes of DTC progression/recurrence

We compared the percentage of DTC progression/recurrence at a mean follow-up of 4.48 years after treatment between the pregnancy (n = 102) and PSM matched non-pregnancy groups (n = 297) in Table 2. There was no evidence of difference between the pregnancy group and the matched non-pregnancy group in growth in the size of existing metastatic foci [2 (2.0 %) vs. 2 (0.7 %); *P* = 0.346], percentage of patients developing new lymph node metastases [4 (3.9 %) vs. 21 (7.1 %); *P* = 0.519], node growth in the contralateral thyroid lobe [4 (3.9 %) vs. 16 (5.4 %); *P* =0.324 ], or biochemical progression [2 (2.0 %) vs. 9 (3.0 %); *P* = 0.583].

**Table 2.** Follow-up outcomes of DTC progression/recurrence in pregnancy and PSM matched non-pregnancy groups

| Outcomes | Pregnancy group<br>(n = 102) | PSM matched<br>non-pregnancy<br>group<br>(n = 297) | P<sup>2</sup> |
| --- | --- | --- | --- |
| **Interval between surgery and follow up (year), mean ± SD** | 5.05 ± 2.04 | 4.29 ± 2.32 | 0.677 |
| **Meeting the treatment goal of TSH suppression, n (%)** | 83 (81.4) | 250 (84.2) | 0.591 |
| **TSH (uIU/ml), median (range)** | 1.08 (0.29-4.17) | 0.61 (0.02-11.70) | 0.795 |
| **Tg (ng/ml), median (range)** | 0.75 (0.04-1.22) | 0.08 (0.04-40.31) | 0.631 |
| **Tg antibody (U/ml), median (range)** | 20.78(15-500) | 15.71 (10-500) | 0.412 |
| **Progression** | 12(11.8) | 47 (15.8) | 0.350 |
| **Structural progression, n (%)** | 10 (9.8) | 38 (12.8)<sup>1</sup> | 0.425 |
| ≥ 3 mm growth in the size of existing metastatic foci | 2 (2.0) | 2(0.7) | 0.346 |
| Development of new lymph node metastases | 4(3.9) | 21 (7.1) | 0.519 |
| ≥ 2 mm growth in the size of existing cancer foci in the contralateral thyroid | 4 (3.9) | 16 (5.4) | 0.324 |
| **Biochemical progression, n (%)** | 2 (2.0) | 9 (3.0) | 0.583 |

**Note:**<sup>1</sup> One patient had two concurrent structural progression, development of new lymph node metastases and ≥ 2 mm growth in the size of existing cancer foci in the contralateral thyroid. <sup>2</sup>T-test of regression based on treatment variables and weighted with matching weights.

### Association of pregnancy with progression risk of DTC

Results from conditional logistic regressions showed no association of pregnancy with progression risk of DTC (*P* > 0.05), after adjusting for the potential confounders of age, tumor size, initial risk stratification, Hashimoto’s thyroiditis, lymph node dissection, the time interval between surgery and follow-up, and achievement of TSH inhibition target (Table 3).

**Table 3.**
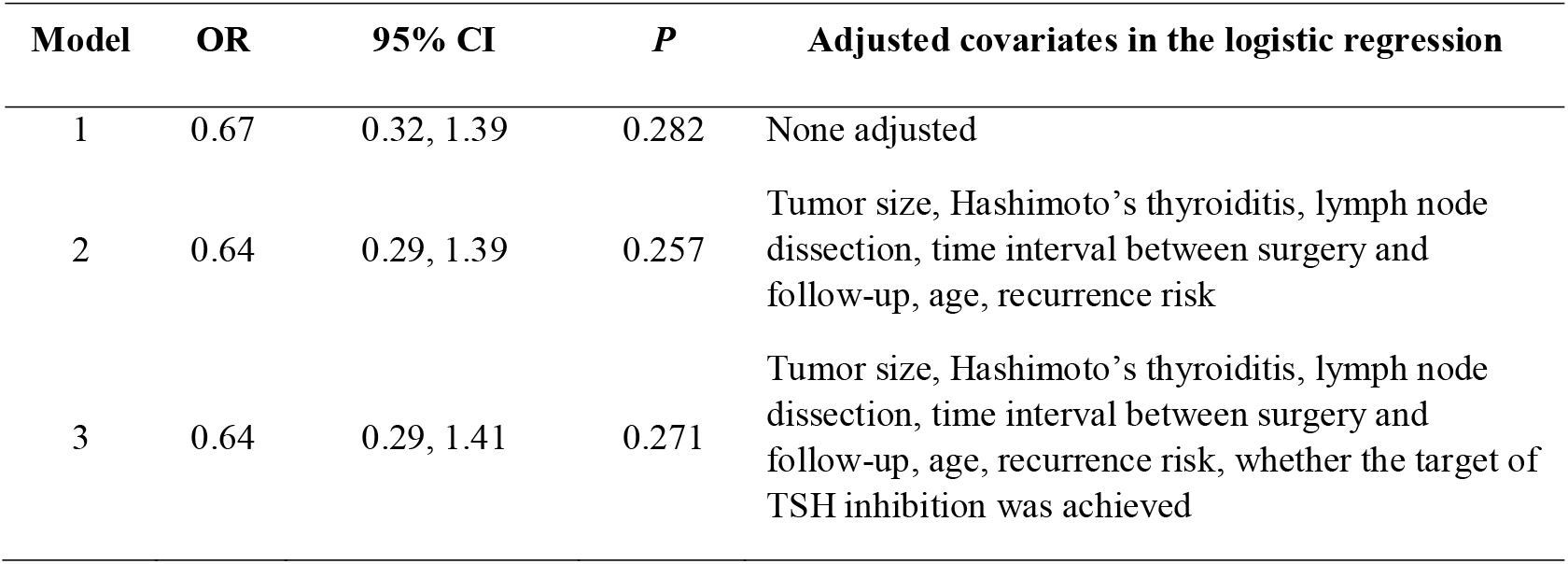
Association outcomes of pregnancy with progression risk of DTC in conditional logistic regression analyses

| **Model** | **OR** | **95% CI** | ***P*** | **Adjusted covariates in the logistic regression** |
| --- | --- | --- | --- | --- |
| 1 | 0.67 | 0.32, 1.39 | 0.282 | None adjusted |
| 2 | 0.64 | 0.29, 1.39 | 0.257 | Tumor size, Hashimoto's thyroiditis, lymph node dissection, time interval between surgery and follow-up, age, recurrence risk |
| 3 | 0.64 | 0.29, 1.41 | 0.271 | Tumor size, Hashimoto's thyroiditis, lymph node dissection, time interval between surgery and follow-up, age, recurrence risk, whether the target of TSH inhibition was achieved |

The pregnancy-progression association observed longer than 4.5 years showed no evidence of difference with that observed shorter than 4.5 years (*P* for interaction was 0.283; Supplementary Table 1). The main results also did not differ by the achievement of the target of TSH inhibition, operation type, or Hashimoto’s thyroiditis (*P*_interaction_ > 0.05, Supplementary Table 1). Results from sensitivity analyses that excluded 11 patients recommended for I^131^ radioactive iodine treatment did not alter the results from the main analyses (Supplementary Table 2).

### Subgroup analyses in the pregnancy group based on the time interval between treatment and pregnancy and pre-pregnancy response to therapy

We further classified the pregnancy patients into 3 subgroups based on the time interval between treatment and pregnancy (< 1 year, 1-2 years, ≥ 2 years) and found that the shorter the time interval, the higher the risk of DTC progression (*P* for trend was 0.043); patients who became pregnant within 1 year after treatment had a higher risk of progression than those became pregnant more than 2 years after treatment [OR: 23.44, 95%CI: 5.08, 108.13; *P* < 0.001] (Table 4).

**Table 4.** Subgroup analysis of progression risk in pregnancy group according to the time interval between pregnancy and surgery

| **Subgroups** | **N** | **Progression,**<br>**n (%)** | **Adjusted**<br>**OR<sup>1</sup>** | **95% CI** | **P** |
| --- | --- | --- | --- | --- | --- |
| More than 2 years | 45 | 2 (4.4) | - | - | - |
| Within 1 to 2 years | 32 | 5 (15.6) | 5.20 | 0.94, 28.67 | 0.058 |
| Within 1 year | 25 | 5 (20.0) | 23.44 | 5.08, 108.13 | <0.001 |

<sup>1</sup>Comared with the first subgroup, adjusted for tumor size, Hashimoto's thyroiditis, lymph node dissection, time interval between surgery and follow-up, age, and recurrence risk.

In the pregnancy group, patients who had a structural incomplete response to therapy before pregnancy had the highest risk of structural and biochemical progression, compared to those having an excellent, indeterminate, or biochemical incomplete response (Supplementary Table 3).

### Evaluation of serum Tg and TSH levels before pregnancy, during pregnancy, and after delivery in the pregnancy group

The mean TSH level during pregnancy in the cases with DTC progression was not substantially higher than the figure of those without DTC progression (Supplementary Figure 1). Among the 8 pregnancy cases with total thyroidectomy and having accurate Tg measurements, only 1 case had elevated Tg levels after delivery (Supplementary Figure 2). For this case, TSH levels increased in early pregnancy and decreased during late pregnancy and postpartum, but Tg levels were maintained at a high degree during the 4-year postpartum period.

## Discussion

We conducted one of the first propensity score-matched retrospective cohort studies to clarify the association of pregnancy with disease progression in patients previously treated for DTC by rigorously comparing the pregnancy group with the matched non-pregnancy group. We observed that pregnancy was not associated with an increased risk of structural or biochemical progression of DTC. This finding remained robust after considering several potential confounders such as Hashimoto’s thyroiditis, treatment with total thyroidectomy or lobectomy, and whether meeting the treatment goal of TSH suppression during pregnancy. However, the time interval between pregnancy and surgery was negatively associate<u>d</u> with the ris<u>k</u> of progression after DTC surgery; patients who became pregnant within 1 year after surgery had a higher risk of progression than those became pregnant more than 2 years after surgery.

Existing studies of the research topic included two types of study design: single-group and controlled-group design. A majority of studies have used a single-group design exclusively focusing on pregnancy patients and their findings remained contradictory ^8,9,20,24,25^. Two of 3 controlled-group studies were limited in the generalizability of their conclusions in terms of sample selection^11,12^ and survival outcomes^12^, as they specifically focused on patients of DTC with distant metastasis ^11,12^ and one study only observed structural progression without the more sensitive measurements of biochemical progression such as thyroglobulin (Tg) levels or Tg antibodies at follow up ^12^. Our study took advantage of the PSM matched controlled-group design to rule out the possibility of a natural disease course leading to progression and to reduce the effect of confounding. Moreover, we carefully evaluated the multiple measurements of TSH levels throughout the preconception, pregnancy, and postpartum period, and found that TSH levels did not differ significantly between patients with DTC progression and those without DTC progression. This observation not only supplemented the previous study which had incomplete data on TSH values during pregnancy ^25^ but also potentially interpreted why the pregnancy was not shown to be associated with DTC progression.

Therefore, the evidence from this study is superior to existing studies in terms of study design, sample selection, outcome measurement, and confounding controlling.

We further stratified patients in the pregnancy group into 4 groups based on distinct response-to-therapy assessments before pregnancy. We found that patients with the structural incomplete outcomes before pregnancy had a higher risk of DTC progression after delivery compared with those with excellent, indeterminate, or biochemical incomplete responses to therapy. This finding, broadly consistent with previous studies ^8,9,20,24,25^, also took a further step. Of note, most DTC patients that have been examined in previous studies were treated with total thyroidectomy (80.6%-100%) ^8,9,20,24,25^, historically regarded as the primary treatment for thyroid cancer. Recent guidelines have recommended lobectomy for most low-risk DTC ^18,26,27^. Thus, our results add to the evidence that pregnancy may not increase the risk of DTC progression even for patients previously treated with lobectomy.

Interestingly, we found that the time interval between treatment and pregnancy was negatively associate<u>d</u> with the ris<u>k</u> of progression after DTC surgery. This finding was consistent with the findings of *Ives A* et al in breast cancer studies^14^. Pregnancy relatively soon after DTC surgery may result in unsatisfactory TSH suppression and changes in the body’s hormone levels, which may contribute to disease progression. However, this finding should be interpreted cautiously due to the small sample size after stratification by time interval. Subsequent research on this issue that confirm our findings will have important practical significance.

Hashimoto’s thyroiditis, the most commonly diagnosed human autoimmune disease, is increasingly prevalent in recent years ^28^. The coexistence between Hashimoto’s thyroiditis and DTC was reported to account for approximately a quarter of patients on average (range: 5%-85%) ^29,30^. Most studies have suggested a potential association of Hashimoto’s thyroiditis with the development and progression of DTC. But relevant evidence is still scarce and contradictory ^13^. To our knowledge, no studies have examined whether Hashimoto’s thyroiditis modified the progression risk of pregnancy-associated DTC. Findings from our study provided initial evidence that this might not be a concerned factor, as the association of pregnancy with DTC progression in patients with Hashimoto’s thyroiditis did not differ from the counterparts.

Generally, we recommend clinicians follow suggestions from ATA guidelines when counseling DTC patients having a future pregnancy plan. In our study population, we found 11.8 % (12 out of 102) of patients previously treated for DTC showed structural or biochemical progression after delivery, and the figure was not higher than that in the matched non-pregnancy group. Notably, we have observed that biochemical progression can exist in a minority of patients for several years after delivery, and whether it is associated with long-term risk of structural progression waits for further investigation.

This study was the largest sample size on this issue to date based on our systematic review^7^. This study, featured in its well-matched control-group design, detailed measurements of both structural and biochemical indicators of progression, and careful control of a multiple of potential confounders by using PSM method, contribute to the study field of pregnancy-associated DTC.

### Limitations

The analysis cohort excluded 21 women (17%) in the pregnancy group whose PS was not within the common range of the two groups, which limited the generalizability of the conclusion to a certain extent. However, this increased the comparability between the pregnant and non-pregnant groups and made results more reliable. Furthermore, even though PSM balanced the baseline covariates as much as possible, it cannot control unmeasured potential confounders^15^. But due to the ethics of randomized controlled trials of the present topic, a well-controlled observational cohort like this study may be one of the feasible solution to answering the study question.

## Conclusions

Among patients previously treated for DTC, the risk of disease progression/recurrence in pregnant women was similar to that in the well-matched, non-pregnant women. Young women previously treated for DTC might not need to worry about their future pregnancy plan in terms of disease progression, but it seems more reasonable to wait an appropriate amount of time before pregnancy.

## Data Availability

All data produced in the present work are contained in the manuscript.

## List of Tables and Figures

**Supplementary Table 1.**
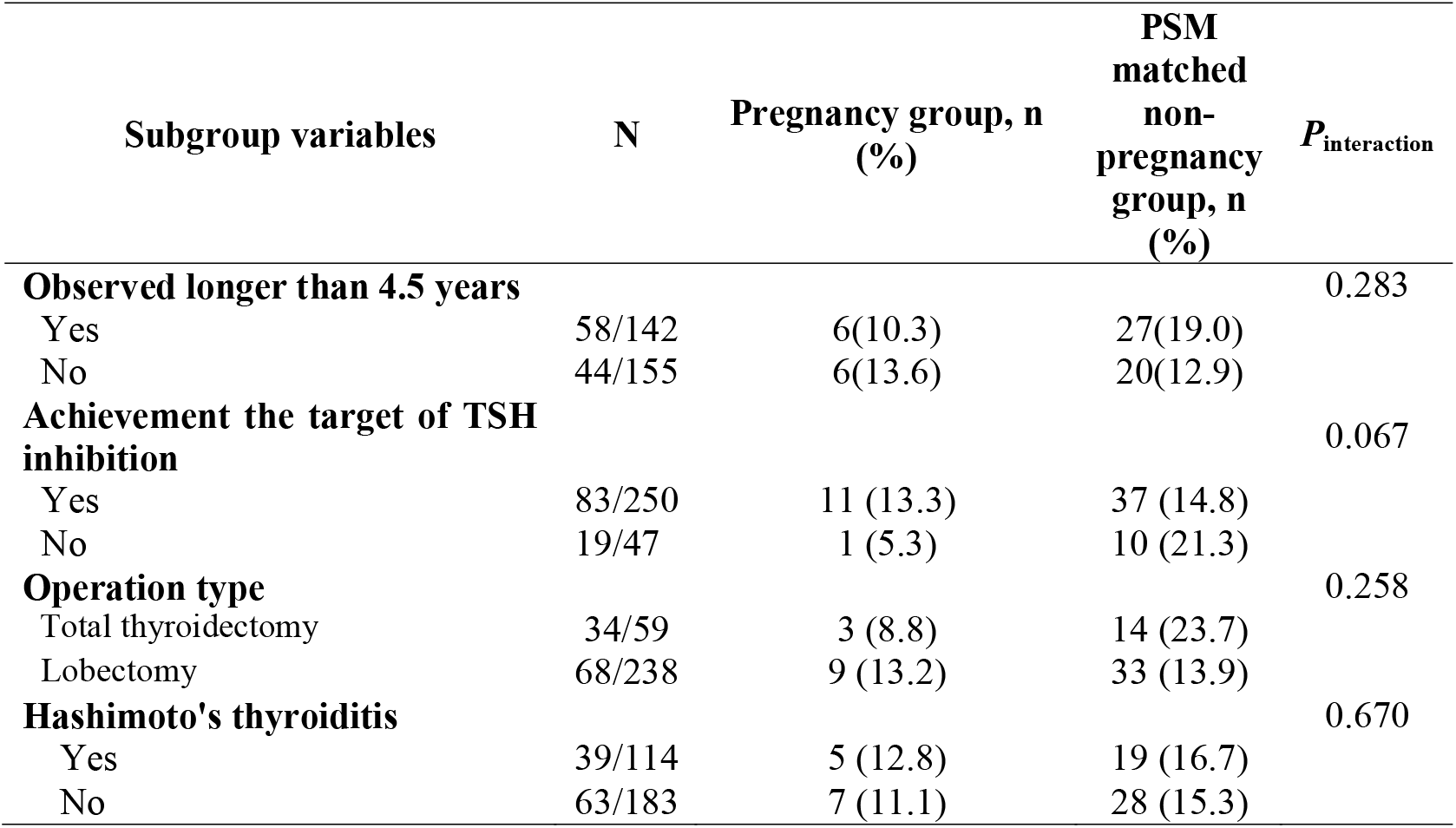
Subgroup analyses of progression risk in pregnancy group and PSM matched non-pregnancy group

**Supplementary Table 2.**
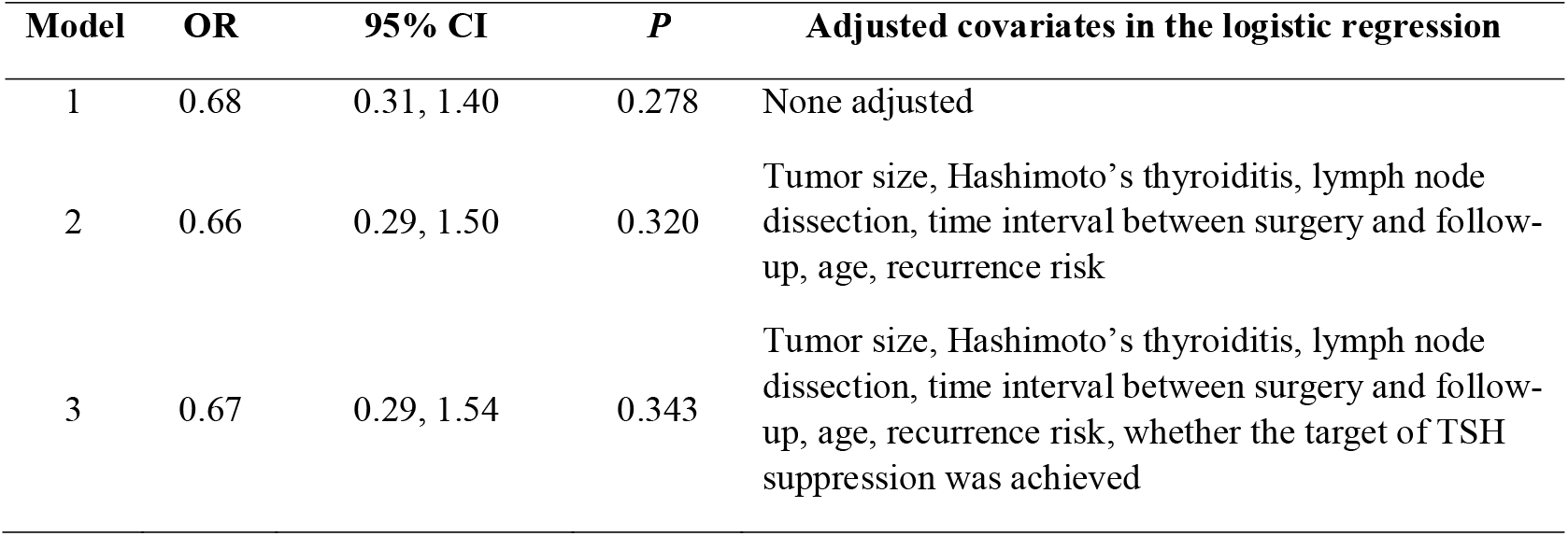
Sensitivity analyses of association of pregnancy with recurrence risk of DTC in conditional logistic regression analyses (excluding patients recommended for I^131^ radioactive iodine treatment)

| Model | OR | 95% CI | *P* | Adjusted covariates in the logistic regression |
| --- | --- | --- | --- | --- |
| 1 | 0.68 | 0.31, 1.40 | 0.278 | None adjusted |
| 2 | 0.66 | 0.29, 1.50 | 0.320 | Tumor size, Hashimoto's thyroiditis, lymph node dissection, time interval between surgery and follow-up, age, recurrence risk |
| 3 | 0.67 | 0.29, 1.54 | 0.343 | Tumor size, Hashimoto's thyroiditis, lymph node dissection, time interval between surgery and follow-up, age, recurrence risk, whether the target of TSH suppression was achieved |

**Supplementary Table 3.** Analyses of the structural or biochemical progression after delivery based on pre-pregnancy response-to-therapy assessments in the pregnancy group

| **Response to therapy** | **No. of cases** | **Structural progression, n (%)** | **Biochemical progression, n (%)** |
| --- | --- | --- | --- |
| Excellent | 51 | 4 (7.8) | 1 (2.0) |
| Indeterminate | 19 | 1 (5.3) | 1(5.3) |
| Biochemical incomplete | 5 | 0 | 0 |
| Structural incomplete | 7 | 2 (28.6) | 0 |
| Missing | 20 | 3 (15.0) | 0 |

**Supplementary Figure 1.**
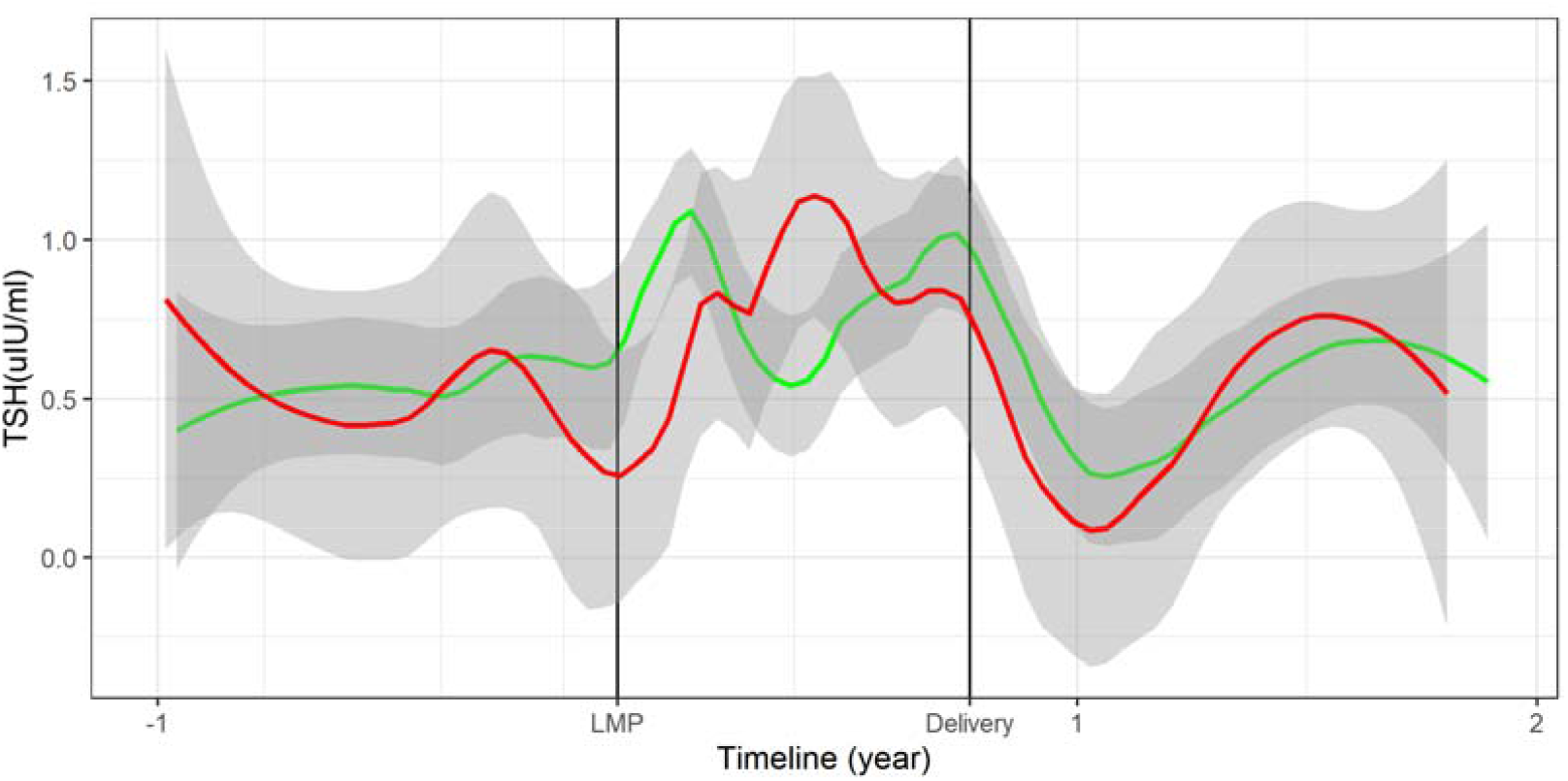
Comparison of TSH levels between patients with DTC progression and those without progression in the pregnancy group (Notes: red line: patients with DTC progression; green line: patients without DTC progression)

**Supplementary Figure 2.**
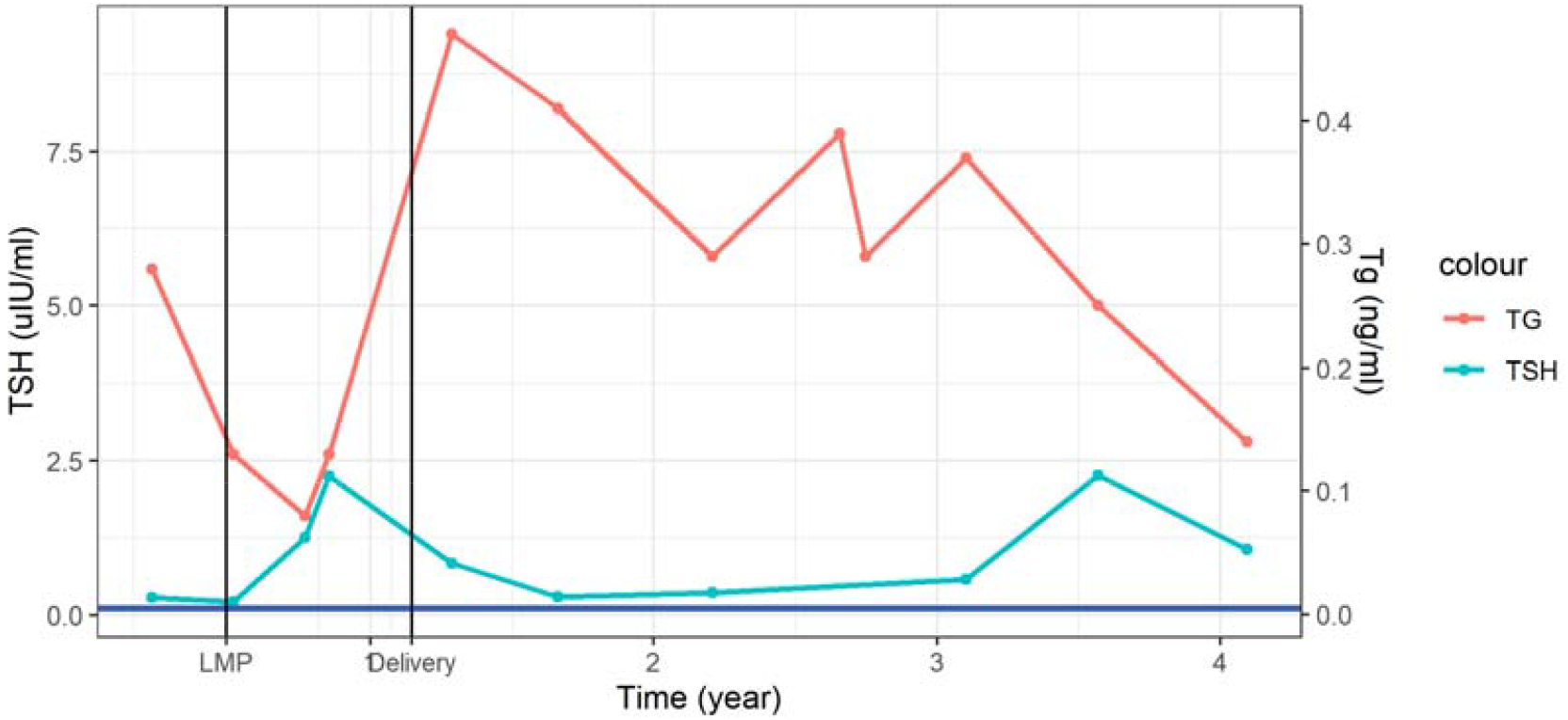
Change of TSH and Tg levels during pre-pregnancy, pregnancy, and postpartum period in 1 case with biochemical progression

## Notes

### Competing Interest Statement

The authors have declared no competing interest.

### Funding Statement

The study was funded by the National Natural Science Foundation of China (81903433; 81672862) and the Fundamental Research Funds for the Central Universities (BMU2021YJ030).

### Author Declarations

This study was approved by the Medical Research Ethics Committee of Peking University Third Hospital (No. IRB00006761-M2022721).

